# SalivaDirect: A simplified and flexible platform to enhance SARS-CoV-2 testing capacity

**DOI:** 10.1101/2020.08.03.20167791

**Authors:** Chantal B.F. Vogels, Anne E. Watkins, Christina A. Harden, Doug E. Brackney, Jared Shafer, Jianhui Wang, César Caraballo, Chaney C. Kalinich, Isabel M. Ott, Joseph R. Fauver, Eriko Kudo, Peiwen Lu, Arvind Venkataraman, Maria Tokuyama, Adam J. Moore, M. Catherine Muenker, Arnau Casanovas-Massana, John Fournier, Santos Bermejo, Melissa Campbell, Rupak Datta, Allison Nelson, Yale IMPACT Research Team, Charles S. Dela Cruz, Albert I. Ko, Akiko Iwasaki, Harlan M. Krumholz, JD Matheus, Pei Hui, Chen Liu, Shelli F. Farhadian, Robby Sikka, Anne L. Wyllie, Nathan D. Grubaugh

## Abstract

Current bottlenecks for improving accessibility and scalability of SARS-CoV-2 testing include diagnostic assay costs, complexity, and supply chain shortages. To resolve these issues, we developed SalivaDirect, which received Emergency Use Authorization (EUA) from the U.S. Food and Drug Administration on August 15th, 2020. The critical component of our approach is to use saliva instead of respiratory swabs, which enables non-invasive frequent sampling and reduces the need for trained healthcare professionals during collection. Furthermore, we simplified our diagnostic test by ***(1)*** not requiring nucleic acid preservatives at sample collection, ***(2)*** replacing nucleic acid extraction with a simple proteinase K and heat treatment step, and ***(3)*** testing specimens with a dualplex quantitative reverse transcription PCR (RT-qPCR) assay. We validated SalivaDirect with reagents and instruments from multiple vendors to minimize the risk for supply chain issues. Regardless of our tested combination of reagents and instruments from different vendors, we found that SalivaDirect is highly sensitive with a limit of detection of 6-12 SARS-CoV-2 copies/μL. When comparing SalivaDirect to paired nasopharyngeal swabs using the authorized ThermoFisher Scientific TaqPath COVID-19 combo kit, we found high agreement in testing outcomes (>94%). In partnership with the National Basketball Association (NBA) and Players Association, we conducted a large-scale *(n* = 3,779) SalivaDirect usability study and comparison to standard nasal/oral tests for asymptomatic and presymptomatic SARS-CoV-2 detection. From this cohort of healthy NBA players, staff, and contractors, we found that 99.7% of samples were valid using our saliva collection techniques and a 89.5% positive and >99.9% negative test agreement to swabs, demonstrating that saliva is a valid and noninvasive alternative to swabs for large-scale SARS-CoV-2 testing. SalivaDirect is a flexible and inexpensive ($1.21-$4.39/sample in reagent costs) option to help improve SARS-CoV-2 testing capacity. Register to become a designated laboratory to use SalivaDirect under our FDA EUA on our website: publichealth.yale.edu/salivadirect/.

## Introduction

SARS-CoV-2, a novel beta-coronavirus, emerged in late 2019 in Wuhan, China, and the subsequent COVID-19 pandemic rapidly followed (Wu et al., 2020; Zhou et al., 2020). In many parts of the world, including the United States, COVID-19 cases continue to rise (Dong et al., 2020; World Health Organization, 2020). The implementation of mass testing efforts followed by contact tracing will be necessary to quell the pandemic. Routine state-level screening and surveillance of healthy individuals is particularly important for safe re-opening of the economy and schools and can minimize the risk of relapsing local outbreaks. However, the scalability and availability of currently authorized assays for SARS-CoV-2 diagnostic testing are still limited, and large-scale application is hampered by worldwide supply chain issues (Patel et al., 2020). To overcome these challenges, mass testing efforts must be ***(1)*** safe, both at the point of specimen collection and specimen processing, ***(2)*** affordable, ***(3)*** flexible, without the need for specific reagents or instrumentation from specific vendors, ***(4)*** adaptable to high-throughput workflows, and ***(5)*** amenable to quick turn-around times. While several different types of diagnostic assays have been recently authorized for emergency use by the U.S. Food and Drug Administration (FDA) such as RT-qPCR, LAMP, CRISPR, and sequencing-based assays, alternatives are still needed for large-scale testing efforts (U.S. Food & Drug Administration, 2020a).

Based on established diagnostic practices for other respiratory infections, the nasopharyngeal swab was initially adopted as the preferred sampling technique for SARS-CoV-2. However, we and others have shown that saliva can serve as an alternative upper respiratory tract specimen type for SARS-CoV-2 detection (Azzi et al., 2020; Iwasaki et al., 2020; To et al., 2020; Wyllie et al., 2020). This is significant as saliva offers a number of advantages over nasopharyngeal swabs when considering the aforementioned criteria for mass testing efforts. Specifically, saliva does not require a certified swab and collection receptacle and does not necessarily have to be obtained by a skilled healthcare provider, both of which increase diagnostic-associated costs. Nasopharyngeal sampling requires a swab being inserted into the back of the nares, which can cause irritation that could promote sneezing and coughing. Thus, the non-invasive collection of saliva is safer as it protects healthcare workers from being inadvertently exposed to potentially infectious droplets. In addition to being more affordable and safer, collection of nasopharyngeal swabs has been associated with variable, inconsistent, and false-negative test results due to the technical difficulties of taking a proper swab (Kinloch et al., 2020; Li et al., 2020; Winichakoon et al., 2020; Wölfel et al., 2020; Wyllie et al., 2020; Zou et al., 2020).

To increase testing capacity for large-scale screening efforts, we developed SalivaDirect, a saliva based, nucleic acid extraction-free, dualplex RT-qPCR method for SARS-CoV-2 detection. Our approach can be broadly implemented as it does not require expensive saliva collection tubes containing preservatives (Ott et al., 2020b), and does not require specialized reagents or equipment for nucleic acid extraction. We validated SalivaDirect for use with products from multiple vendors. Thus, the simplicity and flexibility of SalivaDirect mean that it will not be as affected by supply chain bottlenecks as some other assays that rely on swabs and/or nucleic acid extraction. We show that SalivaDirect has a low limit of detection (6-12 copies/μL) and yields highly concordant results as compared to currently validated RT-qPCR assays. The unique features of SalivaDirect is that it is non-invasive, less expensive ($1.21-$4.39/sample in reagents), and is validated for use with reagents and instruments from multiple vendors. Through our partnerships with the National Basketball Association (NBA) and the National Basketball Players Association (NBPA), we conducted a large usability study of SalivaDirect and comparison to standard RT-qPCR testing of paired anterior nare/oropharyngeal (AN/OP) swabs for asymptomatic/presymptomatic detection of SARS-CoV-2. Our results demonstrate how our specialized protocols for saliva collection produces mostly valid specimens for testing (99.7%) and the specificity of SalivaDirect leads to very few false positive results (0.03-0.05%), showcasing to SalivaDirect - and saliva testing in general - can be used to enhance large-scale testing of symptomatic and asymptomatic individuals. The data presented here were used to support our Emergency Use Authorization (EUA) for SalivaDirect granted by the FDA on August 15th, 2020 (U.S. Food & Drug Administration, 2020b).

## Results

### Development of a simplified SARS-CoV-2 molecular diagnostic framework

To reduce the cost, time, and effort for SARS-CoV-2 detection, we developed SalivaDirect (publichealth.yale.edu/salivadirect/), a simplified and flexible saliva-based platform. SalivaDirect consists of three steps: ***(1)*** collecting saliva without preservative buffers, ***(2)*** proteinase K treatment and heat inactivation in place of nucleic acid extraction, and ***(3)*** dualplex RT-qPCR SARS-CoV-2 detection (**Fig. 1a**).

**Fig. 1:**
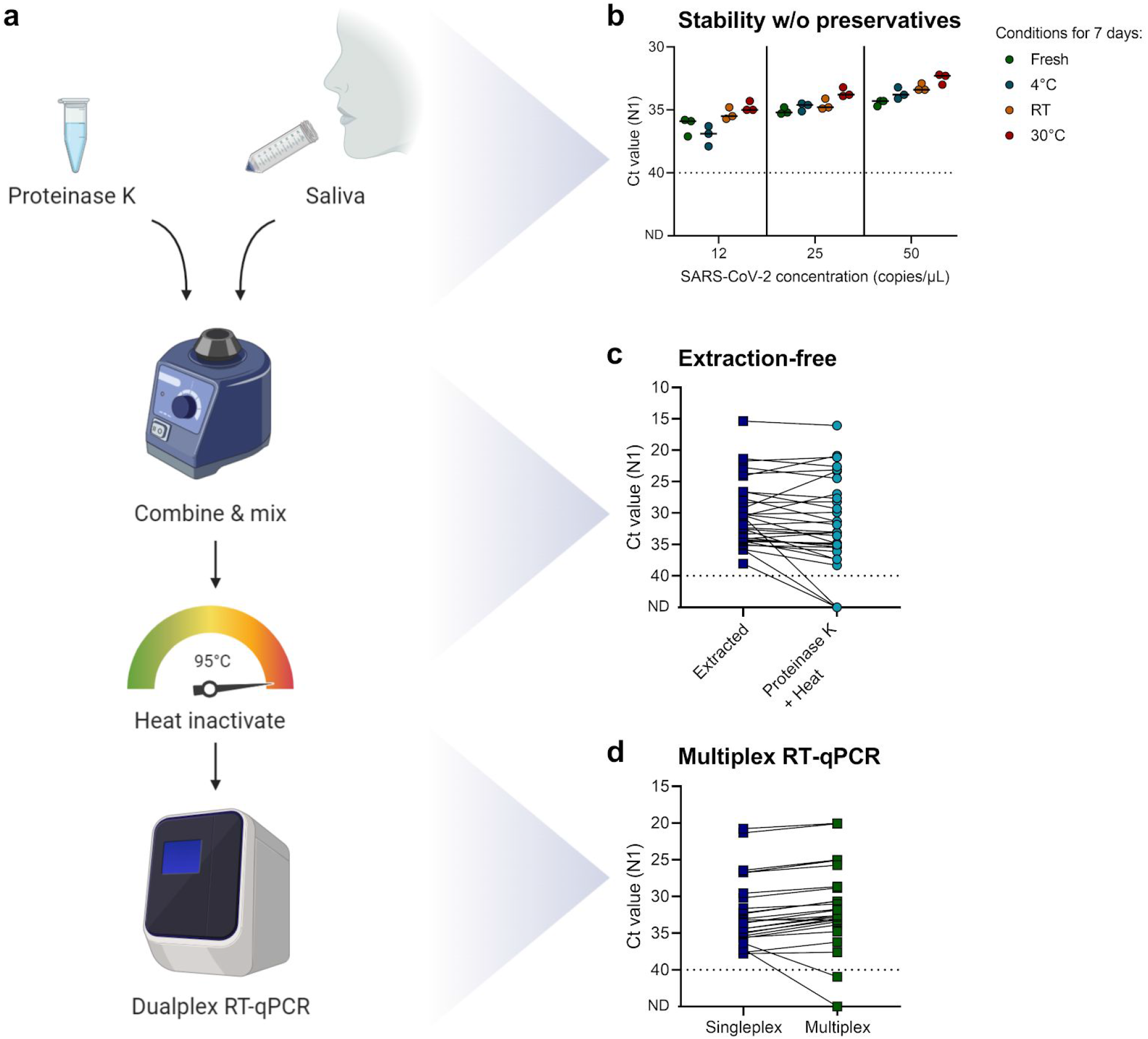
SalivaDirect is a simplified method for SARS-CoV-2 detection. (**a**) Schematic overview of SalivaDirect workflow depicting the main steps of mixing saliva with proteinase K, heat inactivation, and dualplex RT-qPCR testing. Figure created with Biorender.com. (**b**) SARS-CoV-2 is stable in saliva for at least 7 days at 4°C, room temperature (RT, ~19°C), and 30°C without addition of stabilizing buffers. Spiked-in saliva samples of low virus concentrations (12, 25, and 50 SARS-CoV-2 copies/μL) were kept at the indicated temperature for 7 days, and then tested with SalivaDirect. N1 cycle threshold (Ct) values were lower when kept for 7 days at 30°C as compared to fresh specimens (Kruskal-Wallis; *P* = 0.03). Horizontal bars indicate the median. (**c**) Comparing Ct values for saliva treated with proteinase K and heat as compared to nucleic extraction yields higher N1 Ct values without extraction (Wilcoxon; *P* < 0.01). (**d**) Testing extracted nucleic acid from saliva with the N1 primer-probe set (singleplex) as compared to a multiplex assay showed stronger N1 detection in multiplex (Wilcoxon; *P* < 0.01). The dotted line in panels b, c, d indicates the limit of detection. Data used to make this figure can be found in **Source Data Fig. 1**.

### Stability of SARS-CoV-2 detection in saliva without preservatives

Several protocols imply that stabilizing buffers *(e.g*. TBE, TE, or PBS) and additives *(e.g*. Triton-X-100, Tween 20, or NP-40) are required to preserve the detection of SARS-CoV-2 RNA in saliva specimens, while other studies suggest that these buffers are not required and may even inhibit RT-qPCR (Griesemer et al., 2020; Ott et al., 2020b; Ranoa et al., 2020). To determine the stability of SARS-CoV-2 RNA detection using SalivaDirect, we stored saliva specimens for 7 days at 4°C, room temperature, or 30°C without the addition of preservatives. We quantified the virus copies from a positive saliva specimen and spiked-in different concentrations of 12, 25, and 50 SARS-CoV-2 copies/μL into negative saliva collected from healthcare workers (Wyllie et al., 2020). After 7 days, we tested the spiked-in saliva specimens with SalivaDirect and compared results to “fresh” samples. We found that SARS-CoV-2 detection was stable in saliva for at least 7 days at each of the three thermal conditions (**Fig. 1b**). Surprisingly, we even detected significantly lower N1 Ct values *(e.g*. better detection) when saliva was kept for 7 days at 30°C as compared to fresh specimens (median difference across concentrations of 1.4 Ct, *P* = 0.03; **Fig 1b**). In contrast, we found that Ct values for human *RNase P* (RP) were significantly higher after 7 days at RT (median difference of 3.8 Ct, *P* < 0.01) or 30°C (median difference of 5.0 Ct, *P* < 0.001) as compared to fresh specimens, which suggests that the human RNA degraded over time (**Supplementary Fig. 1**). Thus, our data suggest that SARS-CoV-2 RNA is stable in saliva without preservatives for at least 7 days when stored at temperatures of up to 30°C.

### Nucleic acid extraction-free PCR detection of SARS-CoV-2

Nucleic acid extraction is included in most authorized PCR diagnostic assays to detect SARS-CoV-2 RNA by RT-qPCR. However, nucleic acid extraction is relatively expensive, time-consuming, and subject to supply chain bottlenecks which limit the scalability of testing which is critical for safe reopenings. Previous studies have shown that the nucleic acid extraction step can be omitted with a relatively small impact on analytical sensitivity (Arizti-Sanz et al., 2020; Mallmann et al., 2020; Marzinotto et al., 2020; Ranoa et al., 2020; Smyrlaki et al., 2020). Therefore, we explored the potential of proteinase K and heat as an affordable, fast, and easy alternative to nucleic acid extraction. We used the modified CDC assay (Vogels et al., 2020b) to compare RT-qPCR detection of SARS-CoV-2 in saliva specimens processed with nucleic acid extraction or by simply mixing the specimen with proteinase K followed by heat inactivation (**Fig. 1c**). As compared to nucleic acid extraction, our data show that our extraction-free approach minimally decreases detection (median N1 Ct increase = 1.8 Ct; *P* < 0.01). The reduction in detection that we observed is equivalent to what we would expect from omitting the ~4-fold concentration step that occurs during nucleic acid extraction. Our findings demonstrate that proteinase K and heat can be used as an alternative to nucleic acid extraction with only a minor loss in sensitivity.

### Dualplex PCR detection of SARS-CoV-2 and a human control gene

Our final modification to improve the scalability of SARS-CoV-2 diagnostic assays was to increase the high-throughput testing potential of the RT-qPCR step. We previously found that the US CDC primer-probe sets are among the most sensitive and reliable for SARS-CoV-2 detection (Vogels et al., 2020b). The CDC assay consists of three separate reactions targeting two regions of the SARS-CoV-2 nucleocapsid (N1 and N2) and a human *RNase P* (RP) control (Lu et al., 2020). We previously modified the CDC assay by multiplexing the three primer-probe sets, thereby reducing the number of tests from three to one, without a significant impact on its sensitivity (Kudo et al., 2020). When testing the multiplex RT-qPCR assay on saliva treated with proteinase K and heat, however, we were not able to detect consistent results for the N2 primer-probe set, nor for the Sarbeco-E (E) or HKU-ORF1 (ORF1) primer-probe sets with HEX fluorophores (**Supplementary Table 1**). By comparing the N1 and N2 primer-probe sets for 613 clinical samples COVID-19 patients and infected healthcare workers, we found that the median N1 Ct values were 1.2 Ct lower as compared to N2 (**Supplementary Fig. 2**), indicating more consistent and significantly stronger detection (P < 0.001). Therefore, to further simplify the RT-qPCR assay we developed a dualplex RT-qPCR assay based on N1 and RP, and modified the fluorophore (Cy5, ATTO647, or Quasar670 instead of FAM) on the RP probe. When comparing the modified singleplex CDC assay with the dualplex assay on extracted nucleic acid, median N1 Ct values were 0.9 Ct lower when tested in multiplex (P < 0.01; **Fig. 1d**). Thus, SalivaDirect allows for a reduction in the number of RT-qPCR reactions to one reaction per sample.

### Lower limit of detection using reagents and equipment from multiple vendors

Tests that depend on specific reagents from single vendors leave them vulnerable to supply chain shortages, as happened throughout the COVID-19 pandemic. Thus our approach was to validate SalivaDirect using reagents and instruments from multiple vendors to avoid dependence on a single vendor for each step (**Table 1**). In addition to what is shown here, we will continue to amend our SalivaDirect EUA through the use of bridging studies to ensure that it remains free of supply chain issues and to provide cheaper alternatives. A current list of validated products can be found in our updated EUA summary (U.S. Food & Drug Administration, 2020b)

**Table 1:** Validated reagents and instruments for use with SalivaDirect.

| Item | Vendor | Product Name | Catalog number |
| --- | --- | --- | --- |
| Proteinase K | ThermoFisher Scientific | MagMAX Viral/Pathogen Proteinase K | A42363 |
|  | New England Biolabs | Proteinase K, Molecular Biology Grade | P8107S |
|  | AmericanBio | Proteinase K | AB00925-00100 |
| RT-qPCR kit | New England Biolabs | Luna Universal Probe One-Step RT-qPCR Kit | E3006S |
|  |  |  | E3006L |
|  |  |  | E3006X |
|  |  |  | E3006E |
|  | Bio-Rad | Reliance One-Step Multiplex RT-qPCR Supermix | 12010176 |
|  |  |  | 12010220 |
|  |  |  | 12010221 |
|  | ThermoFisher Scientific | TaqPath 1-Step RT-qPCR Master Mix, GC | A15299 |
|  |  |  | A15300 |
| RT-qPCR instrument | Bio-Rad | CFX96 Touch Real-Time PCR Detection System |  |
|  | ThermoFisher Scientific | Applied Biosystems 7500 Fast Real-Time PCR System |  |
|  | ThermoFisher Scientific | Applied Biosystems 7500 Fast Dx Real-Time PCR System |  |

To validate reagents and instruments, we spiked a known concentration of SARS-CoV-2-positive saliva into negative saliva from healthcare workers to prepare a 2-fold dilution series of 400, 200, 100, 50, 25, 12, and 6 virus copies/μL. By testing each concentration in triplicate, we determined the preliminary limit of detection, which was then confirmed by testing another 20 replicates (**Fig. 2**). Treating saliva with proteinase K from three different vendors resulted in a limit of detection of 6 SARS-CoV-2 copies/μL, and suggests that SalivaDirect is not dependent on proteinase K from a specific vendor (**Fig. 2a-c**).

**Fig. 2:**
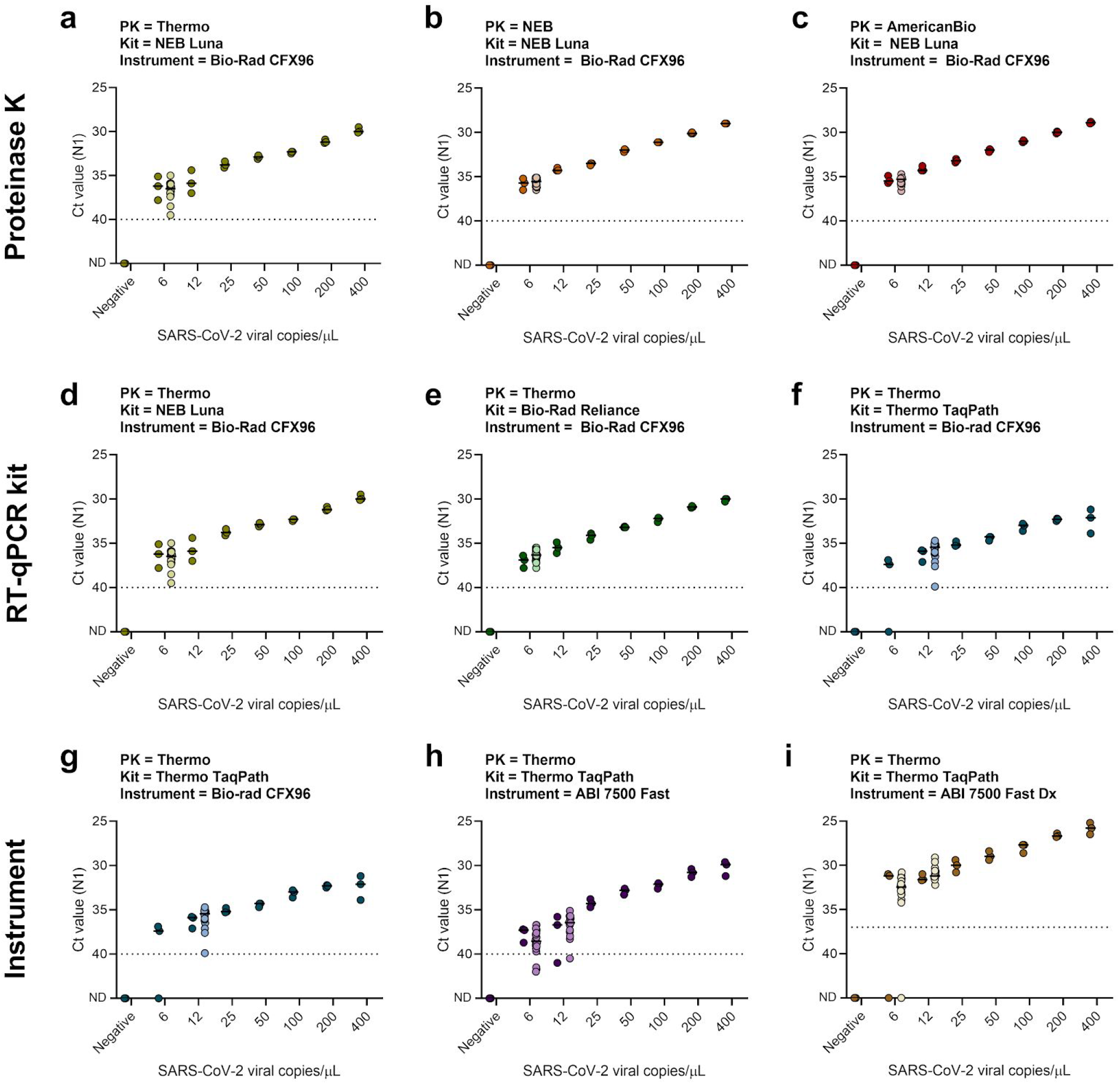
SalivaDirect is validated for use with reagents and instruments from multiple vendors. We determined the lower limit of detection of SalivaDirect with a two-fold dilution series (400, 200, 100, 50, 25, 12, and 6 copies/μL) of positive saliva spiked-in negative saliva. Initially, each concentration and negative saliva were tested in triplicate to determine the preliminary limit of detection (dark-colored dots). The limit of detection was confirmed with 20 additional replicates (light-colored dots) for which 19 out of 20 needed to be detected. Limit of detection when tested with (**a-c**) proteinase K, (**d-f**) RT-qPCR kits, and (**g-i**) RT-qPCR instruments from different vendors, while keeping the other conditions constant. Panels a and d, as well as f and g are duplicates to enable comparisons between the different combinations of reagents or instruments within a single row. Shown are the Ct values for the N1 primer-probe set. The horizontal bars indicate the median and the dotted line indicates the limit of detection. Data used to make this figure can be found in **Source Data Fig. 2**.

Next, we determined the limit of detection by comparing three different RT-qPCR kits obtained from New England Biolabs, Bio-Rad, and ThermoFisher Scientific **(Table 1**). As each kit specifies the use of slightly different PCR cycle times and temperatures, we first sought to standardize these into a “universal” thermocycler program to make it easier to switch between products when needed. Comparing the results from each kit using the manufacturer’s protocol and the universal RT-qPCR program, we found no significant differences in Ct values (Luna: *P* = 0.69, Reliance: *P* = 0.06, TaqPath: *P* = 0.44; **Supplementary Fig. 3**). One additional RT-qPCR kit, Invitrogen EXPRESS One-Step SuperScript qRT-PCR kit, which we tested under their recommended protocol as well as our universal program, did not seem compatible with SalivaDirect and was therefore excluded from our limit of detection experiment. Using the universal thermocycler program with the Bio-rad CFX96 instrument, New England Biolabs (NEB) Luna Universal Probe One-Step kit and Bio-Rad Reliance One-Step Multiplex RT-qPCR Supermix had a lower limit of detection of 6 SARS-CoV-2 copies/μL, whereas the ThermoFisher Scientific TaqPath 1-Step RT-qPCR Master Mix resulted in a slightly higher limit of detection of 12 SARS-CoV-2 copies/μL (**Fig. 2d-f**). Importantly, this indicates that the specific RT-qPCR kit can influence the lower limit of virus detection and not all kits may be suitable for use with SalivaDirect.

Using the qRT-PCR kit with the highest limit of detection, TaqPath 1-Step RT-qPCR Master Mix, we compared the detection across three commonly used RT-qPCR thermocycler instruments: Bio-rad CFX96, Applied Biosystems (ABI) 7500 Fast, ABI 7500 Fast Dx. We found that the Bio-rad CFX96 and ABI 7500 Fast had similar lower limits of detection at 12 SARS-CoV-2 copies/μL, whereas the ABI 7500 Fast Dx had a slightly lower limit of detection of 6 SARS-CoV-2 copies/μL (**Fig. 2g-i**). Interestingly, when determining the preliminary limit of detection for the ABI 7500 Fast Dx, we found that Ct values were on average 4.7 lower than Ct values generated on the ABI 7500 Fast. This suggests a difference in the auto-threshold that the machine sets and therefore, we have increased the positive threshold to 37 Ct for the ABI 7500 Fast Dx to correspond to the positive threshold for the ThermoFisher Scientific TaqPath COVID-19 combo kit using the ABI 7500 Fast Dx. Changing the threshold did not affect the confirmed lower limit of detection of 6 copies/μL for the ABI 7500 Fast Dx. Overall, we found that SalivaDirect has a low limit of detection (6-12 SARS-CoV-2 copies/μL) using reagents and instruments from multiple vendors.

### Sensitivity of SalivaDirect compared to saliva tested using a standard RT-qPCR assay

After determining the lower limit of detection of SalivaDirect, we compared the sensitivity of SARS-CoV-2 detection from saliva using a standard approach - a modified CDC assay with nucleic acid extraction and singleplex RT-qPCR (Vogels et al., 2020b). We found that the median N1 Ct values were 1.2 higher *(e.g*. weaker detection) for SalivaDirect as compared to the modified CDC assay (P < 0.001; **Fig. 3**). Overall, the reduction in analytical sensitivity contributed to a 7.3% (3/41) false negative rate for SalivaDirect, but only for weakly positive samples (all three false negative specimens had N1 Ct values of 35-40 when using the modified CDC assay; **Fig. 3**). Our findings show ~93% positive agreement of SalivaDirect compared to a standard testing approach.

**Fig. 3:**
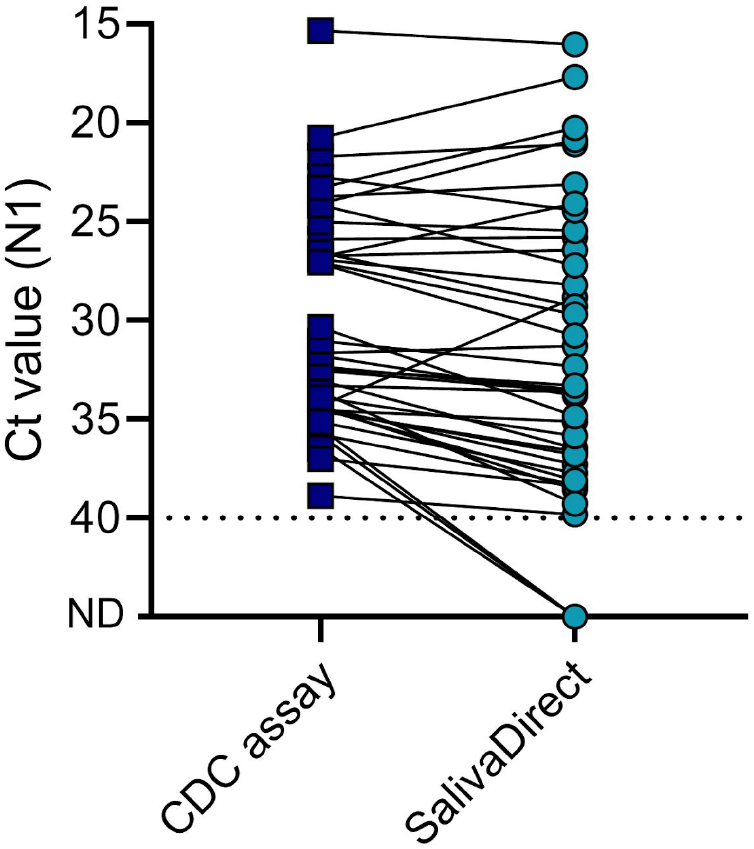
Sensitivity of SalivaDirect is comparable to a standard approach for SARS-CoV-2 detection in saliva. We compared Ct values for N1 between the modified CDC assay (nucleic acid extraction and singleplex RT-qPCR) and SalivaDirect for 41 saliva specimens tested with both methods. Overall, detection of SARS-CoV-2 with SalivaDirect is weaker (median 1.2 Ct, Wilcoxon; *P* < 0.001) than the modified CDC assay, but with a high agreement in outcomes of both tests of (93%). Shown are the Ct values for the N1 primer-probe set and the dotted line indicates the limit of detection. Data used to make this figure can be found in **Source Data Fig. 3**.

### Clinical validation with paired nasopharyngeal swabs and saliva

For consideration of an EUA, the FDA requires clinical validation of any new laboratory developed test by comparing to currently authorized tests. For our validation study, we compared both across tests - SalivaDirect to the authorized ThermoFisher Scientific TaqPath COVID-19 combo kit (Applied Biosystems, 2020) - and across sample types - saliva to nasopharyngeal swabs (**Fig. 4, Tables 2-3**). We collected 37 paired positive and 30 paired negative nasopharyngeal swabs and saliva specimens from inpatients and healthcare workers at the Yale-New Haven Hospital. The ThermoFisher Scientific TaqPath COVID-19 combo kit combines nucleic acid extraction using the MagMax Viral/Pathogen Nucleic Acid Isolation Kit with a multiplex RT-PCR diagnostic assay targeting 3 regions of the SARS-CoV-2 genome on the ABI 7500 Fast Dx instrument. For SalivaDirect, we used the ThermoFisher Scientific proteinase K, ThermoFisher Scientific TaqPath RT-PCR kit, and Bio-Rad CFX96 instrument. We selected the positive and negative pairs based on preliminary results of our modified CDC assay.

**Fig. 4:**
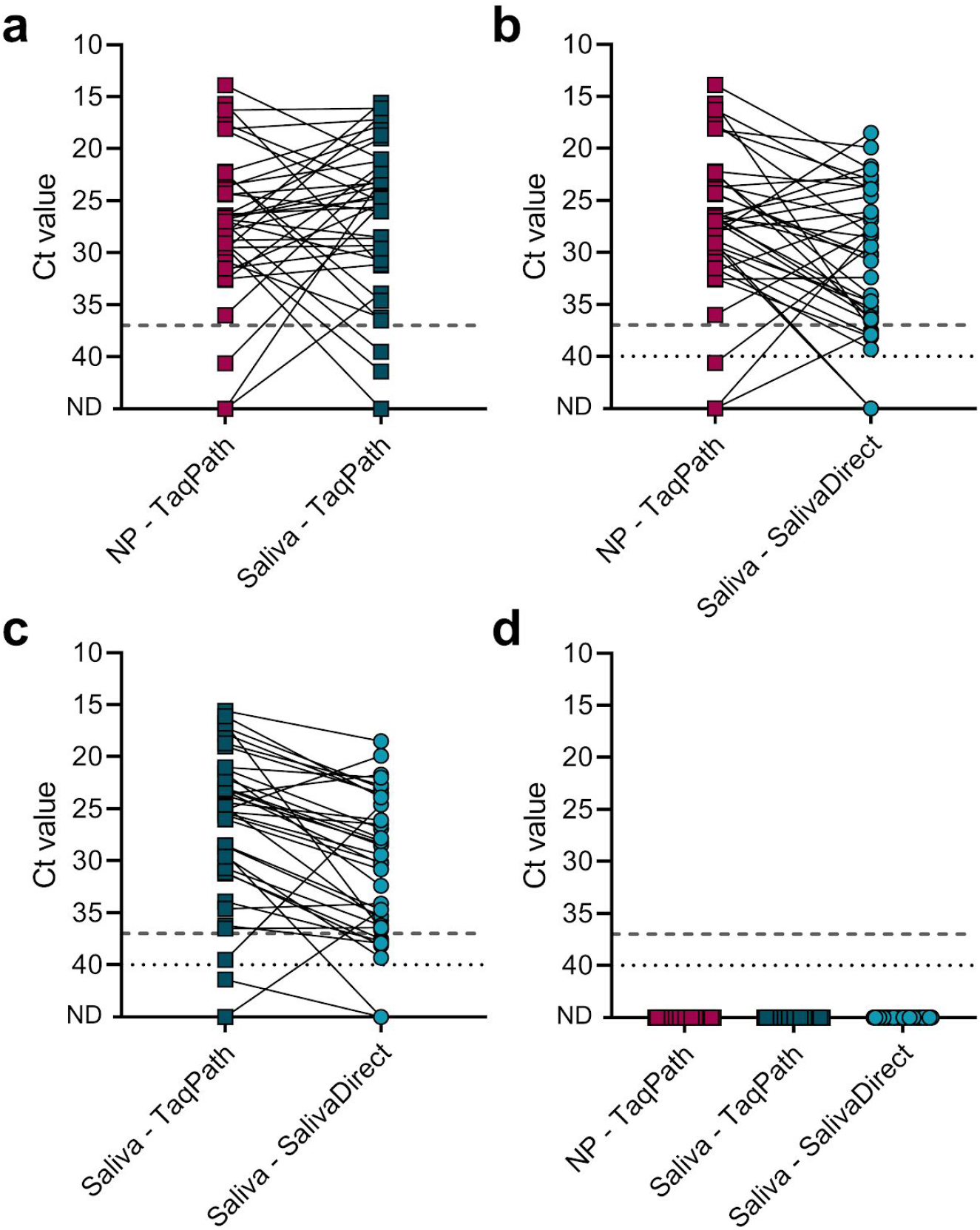
SalivaDirect is highly comparable to standard RT-qPCR tests with nucleic acid extraction from nasopharyngeal swabs and saliva. We selected 37 paired positive and 30 paired negative nasopharyngeal swabs and saliva specimens. Paired samples were collected a maximum 4 days apart. Nasopharyngeal swabs and saliva specimens were tested with the ThermoFisher Scientific TaqPath COVID-19 combo kit and average Ct values for N, S, and ORFlab were compared to N1 Ct values for saliva specimens tested with SalivaDirect. (**a**) Comparison of 37 paired nasopharyngeal swabs and saliva tested with the TaqPath COVID-19 combo kit showed 84% positive agreement, and no significant differences in each of the three virus targets (Wilcoxon; N: *P* = 0.51, S: *P* = 0.72, ORFlab: *P* = 0.39). (**b**) Comparison of nasopharyngeal swabs tested with the TaqPatch COVID-19 combo kit and saliva tested with SalivaDirect showed 94% positive agreement. Median N1 Ct values were 3.3 Ct higher for SalivaDirect (Wilcoxon; *P* < 0.01). (**c**) Comparison of saliva tested with TaqPath COVID-19 combo kit and SalivaDirect again shows that SalivaDirect showed 97% positive agreement. Median N1 Ct values were 5.0 Ct higher for SalivaDirect (Wilcoxon; *P* < 0.001). (**d**) 30 paired nasopharyngeal swabs and saliva specimens tested negative with both the TaqPath COVID-19 combo kit and SalivaDirect. Shown are average Ct values for N, S, and ORF1ab for the TaqPath combo kit and N1 Ct values for SalivaDirect. The dashed line indicates the limit of detection for the TaqPath combo kit (37 Ct) and the dotted line indicates the limit of detection for SalivaDirect (40 Ct). Data used to make this figure can be found in **Source Data Fig. 4**.

**Table 2:**
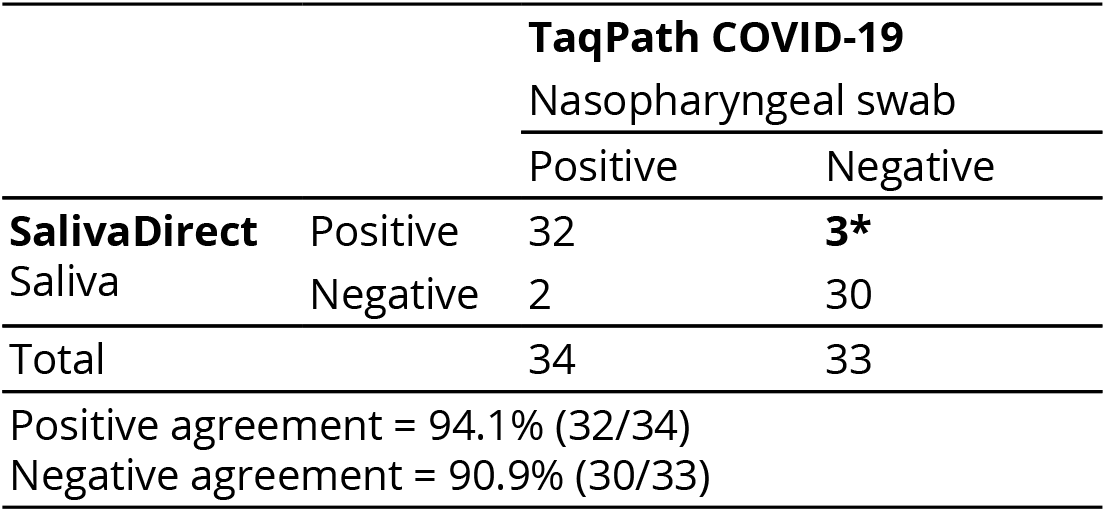
Parallel testing of nasopharyngeal swabs and saliva from inpatients and healthcare workers. Three nasopharyngeal swabs tested negative while previous outcomes of the modified CDC assay indicated that they were weakly positive.

**Table 3:** Parallel testing of saliva from inpatients and healthcare workers using different tests.

|  |  | **TaqPath COVID-19** |  |  |  |
| --- | --- | --- | --- | --- | --- |
|  |  | Saliva |  |  |  |
|  |  | Positive | Inconclusive | Invalid | Negative |
| **SalivaDirect** | Positive | 33 |  | 2 |  |
| Saliva | Negative | 1 | 1 |  | 30 |
| Total |  | 34 | 1 | 2 | 30 |
| Positive agreement = 97.1% (33/34) |  |  |  |  |  |
| Negative agreement = 100% (30/30) |  |  |  |  |  |

First, when we compared nasopharyngeal swabs and saliva specimens when tested with the TaqPath COVID-19 combo kit, we found a positive agreement of 91.2% (**Fig. 4a**). For both sample types there were 3 specimens that tested negative, invalid, or inconclusive while the other sample type tested positive. However, we did not find significant differences in Ct values for the three virus targets between both sample types (P = 0.39-0.72), with the median difference for each of the virus targets <2 Ct. This again confirms that some variation exists between sample types, but that saliva is a valuable alternative (Azzi et al., 2020; Iwasaki et al., 2020; To et al., 2020; Wyllie et al., 2020).

Next, we found a 94% positive agreement with SalivaDirect compared to nasopharyngeal swabs tested with the TaqPath COVID-19 combo kit (**Table 2**). The N1 Ct values were higher using SalivaDirect (median difference of 3.3 Ct; *P* < 0.01; **Fig. 4b**), and the increased Ct values are likely due to a combination of removing the nucleic acid step (**Fig. 1c, Fig. 3**) and using different thermocycler instruments (**Fig. 2**). Out of the 37 nasopharyngeal swabs that were tested with the TaqPath COVID-19 combo kit, three specimens tested negative (**Table 2** and **Fig. 4b**). However, earlier results with the modified CDC assay indicated a (weakly) positive signal, and the paired saliva specimen tested positive with both SalivaDirect and the TaqPath COVID-19 kit. While this is not captured in the percentage of positive agreement, SalivaDirect was able to detect SARS-CoV-2 in saliva of three individuals for which the nasopharyngeal swab tested negative.

When we directly compared the results of SARS-CoV-2 detection from saliva using SalivaDirect and the TaqPath COVID-19 combo kit, we found a high positive (97.1%) as well as negative agreement (100%; **Table 3**). Ct values for N1 were higher when comparing SalivaDirect with the TaqPath COVID-19 combo kit (median difference of 5.0 Ct, *P* < 0.001; **Fig. 4c**) for likely reasons as described above. We intentionally included this comparison to enable a direct comparison of test results based on the same input specimen.

**Table 4:** Parallel testing of anterior nares/oropharyngeal swabs and saliva from NBA players, staff, and contractors.

|  |  | Quest/BioReference |  |
| --- | --- | --- | --- |
|  |  | AN/OP swab |  |
|  |  | Positive | Negative |
| **SalivaDirect**<br>Saliva | Positive | 17 | 2 |
|  | Negative | 2 | 3,746 |
|  | Invalid | 0 | 12 |
| Total |  | 19 | 3,760 |
| Invalid samples = 0.3% (12/3779) |  |  |  |
| Positive agreement = 89.5% (17/19) |  |  |  |
| Negative agreement = 99.9% (3,746/3,748 valid samples) |  |  |  |
| Overall agreement = 99.9% (3763/3767 valid samples) |  |  |  |

Finally, we compared results of negative paired nasopharyngeal swabs and saliva specimens tested with both the TaqPath COVID-19 combo kit and SalivaDirect (**Fig. 4d**). No SARS-CoV-2 was detected in any of the specimens, while we did detect the internal controls. Thus, we did not detect any false positive results with any of the assays.

### Evaluation of off-target amplification

Background amplification or cross-reactivity of primer-probe sets with related human respiratory pathogens can cause false-positive results. Previous *in vitro* evaluations by the CDC showed no cross-reactivity with other human coronaviruses (229E, OC43, NL63, and HKU1), MERS-coronavirus, SARS-coronavirus, and 14 additional human respiratory viruses (Lu et al., 2020). These findings are in accordance with our previous investigation of nine primer-probe sets, including the N1 set, which did not detect any background amplification (Vogels et al., 2020b). To test for possible cross-reactivity of the dualplex RT-qPCR assay, we tested 52 saliva specimens collected from adults in the 2018/2019 and 2019/2020 fall and winter (pre-COVID-19; **Supplemental Fig. 4**). We did not detect off-target amplification or false positives, which is in agreement with previous findings from the CDC (Lu et al., 2020).

### Asymptomatic validation with paired AN/OP swabs and saliva

To conduct effective SARS-CoV-2 screening programs to allow populations to return to school and work, the test must be able to detect asymptomatic and/or presymptomatic cases and have a low false-positive rate. To evaluate SalivaDirect for these uses, we compared 3,779 saliva samples to paired combined AN/OP swabs from healthy NBA players, staff, and contractors (**Table 4**). The saliva samples were sent to the Yale School of Public Health for testing by SalivaDirect and the AN/OP swabs were tested by commercial clinical diagnostic laboratories (Quest Diagnostics or BioReference Laboratories) using standard qRT-PCR assays.

There are concerns with using saliva as a testing specimen as accidental collection of sputum and/or remnants from food or drinks can interfere with sample processing or inhibit PCR. For example, in a study with 124 symptomatic outpatients without optimized saliva collection protocols, approximately a third of the samples were difficult to pipet (Landry et al., 2020). In our NBA cohort study, we developed explicit instructions on how to collect true saliva (Ott et al., 2020a; Vogels et al., 2020a), used a saliva collection aid (a straw that fits into the collection tube) to promote providing true saliva and to minimize potential aerosolization, and added proteinase K as the first laboratory step to help degrade any mucus. As a result, all 3,779 saliva samples collected from the cohort could be tested by SalivaDirect. Furthermore, the use of the RP human control gene in the dualplex PCR helps to determine if there are inhibitors in the specimens. We found that 12 of the 3,779 saliva samples had RP values above a Ct of 35, the threshold for an invalid sample (**Table 4**; **Supplementary Fig. 5**). Thus 0.3% of the saliva samples collected from our cohort were invalid by PCR. Overall, we had a high rate of success for testing saliva by SalivaDirect.

During the study, 19 AN/OP swabs were positive for SARS-CoV-2, and 17 of those were also positive from saliva tested by SalivaDirect (89.5% positive agreement; **Table 4**). Out of the 19 AN/OP swabs that tested positive by commercial clinical diagnostic laboratories, we received ten for comparative testing in our lab with the modified multiplex CDC assay (**Supplementary Fig. 6**). When comparing the Ct values for the paired AN/OP swabs and saliva we found no significant differences between both sample types (P = 0.91; **Supplementary Fig. 6**). Upon retesting with the modified multiplex CDC assay, we found that two AN/OP swabs tested below our positive threshold, indicating that these samples may have been weakly positive or false-positive from the commercial labs. Paired saliva of one of these two swab specimens also tested negative with SalivaDirect. Thus, the true sensitivity of SalivaDirect to AN/OP swabs by standard qRT-PCR for asymptomatic/presymptomatic detection may be higher than 90%; however, a larger positive sample size is needed to further evaluate.

Out of the total number of 3,748 valid samples that tested negative by AN/OP swabs, 3,646 were also identified as negative by SalivaDirect resulting in a negative agreement of 99.9%. For one of the samples that was negative by AN/OP swabs but positive by SalivaDirect, the subsequent saliva and AN/OP swabs from the same individual tested positive, suggesting that the previous SalivaDirect result was a true positive. For the other incongruent result (AN/OP negative, SalivaDirect positive), however, subsequent saliva and AN/OP swabs tested negative, suggesting that the previous SalivaDirect result was a false-positive. Thus, our data indicate that the false-positive rate for SalivaDirect is between 1-2 per 3,748 (0.03-0.05%) samples tested.

### Supply costs for SalivaDirect testing

We aimed to develop a simplified testing method that is not dependent on commercialized kits which may be subject to supply chain issues. Therefore, we reduced the number of steps and initially validated SalivaDirect with reagents and instruments from three different vendors. By doing so we have reduced the cost per sample to a minimum of $1.21, if saliva is collected without a saliva collection aid, and a maximum of $4.39 when using a saliva collection aid (**Table 5**). These cost estimates are based on list prices; therefore the actual costs may be lower. Additional reagents and instruments can be validated by performing a bridging study to show an equal limit of detection and can be submitted to the FDA as an amendment to the authorized EUA. Thus, SalivaDirect provides a relatively inexpensive alternative to current RT-qPCR-based assays.

**Table 5:**
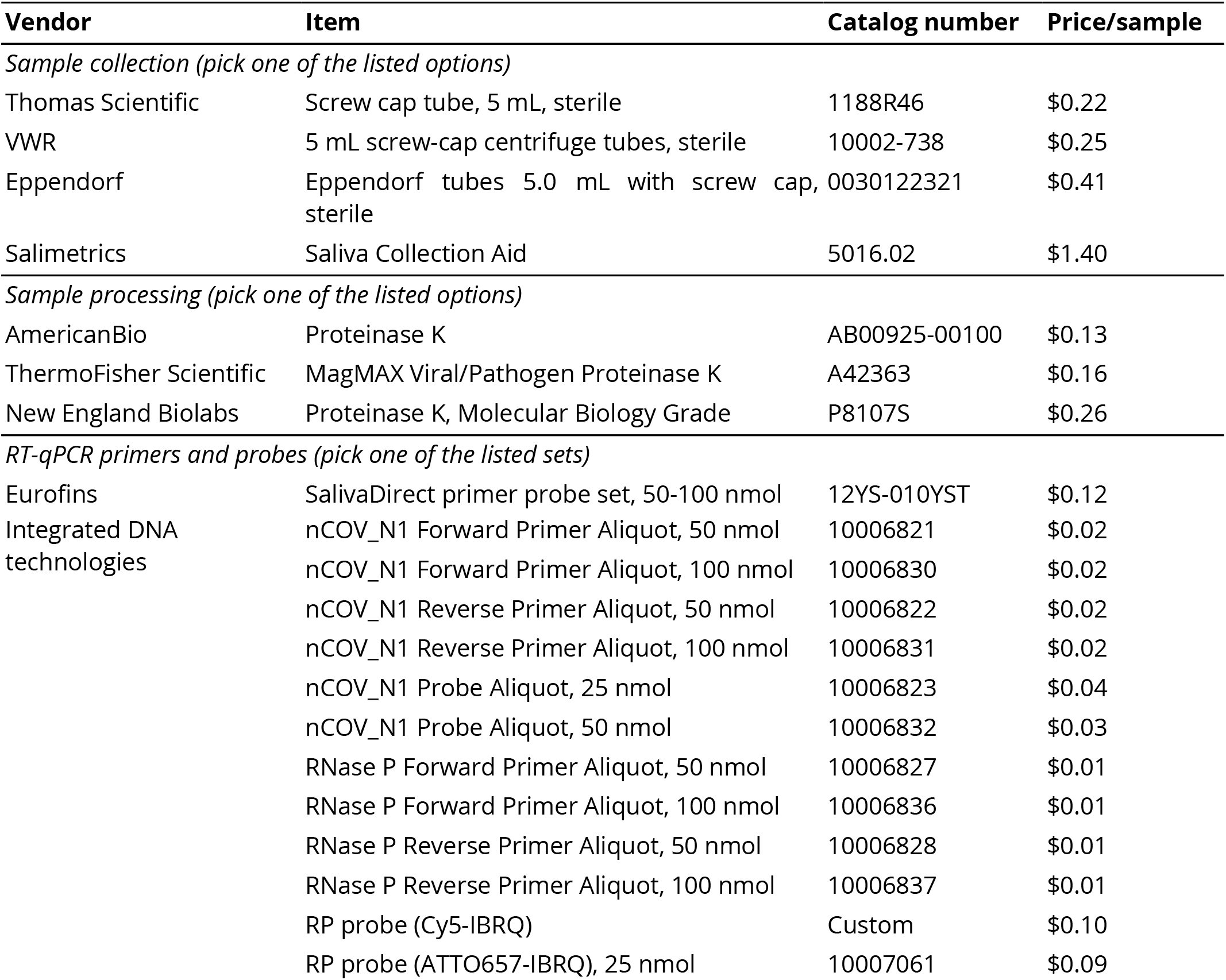
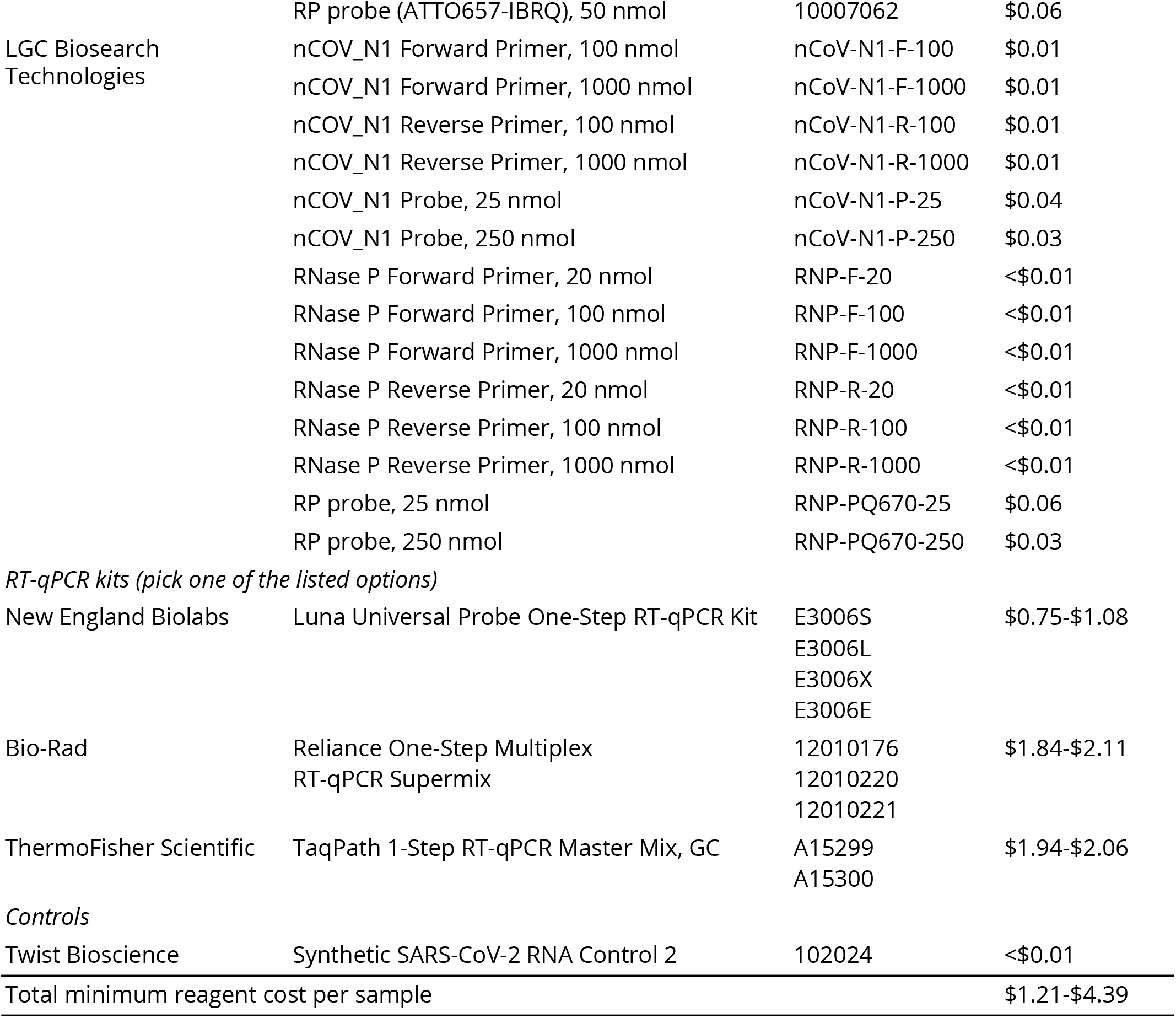
SalivaDirect is a relatively inexpensive method for SARS-CoV-2 diagnostic testing. The price per sample is calculated based on prices listed on the vendor websites and does not include additional costs for general laboratory consumables such as pipet tips or required equipment and instruments such as pipette and RT-qPCR instruments.

## Discussion

### SalivaDirect is a simplified and flexible platform

We developed SalivaDirect to adapt to the needs and budgets of heterogeneous SARS-CoV-2 surveillance systems. Testing saliva as an alternative to invasive swabs allows for safe and easy specimen collection. Furthermore, high-throughput testing can be maximized without the need for expensive saliva collection tubes with stabilizing reagents and nucleic acid extraction kits, and a reduction in RT-qPCR reagents needed per specimen. We validated SalivaDirect with multiple reagents and instruments from different vendors to provide alternative options to minimize bottlenecks associated with supply chain issues. Furthermore, we demonstrated its effectiveness for repeated screening of a relatively young and healthy population from the NBA. Our results from this large cohort indicate a low rate of invalid (0.3%) and false positive (0.03-0.05%) samples when compared to AN/OP swabs tested by commercial clinical diagnostic laboratories, showcasing how our saliva collection and testing protocols are conducive for large-scale asymptomatic SARS-CoV-2 screening. Uniquely, the US FDA authorized us to designate other high-complexity Clinical Laboratory Improvement Amendments (CLIA) certified laboratories to use SalivaDirect, allowing our flexible and simplified framework to help increase the capacity of existing laboratory infrastructure for SARS-CoV-2 testing.

To better adapt to interested laboratories or expand product availability, additional reagents and instruments can be added to our SalivaDirect FDA EUA. This can be done by performing bridging studies to establish equivalent performance between parallel testing of saliva specimens with new and previously validated components (U.S. Food & Drug Administration, 2020c). The FDA recommends testing 3-fold serial dilutions of SARS-CoV-2 spiked saliva specimens in a pooled negative saliva matrix in triplicate, until a hit rate of <100% is reached. Both tests can be considered to have equivalent performance if the resultant limit of detection is the same *(e.g*. <3x limit of detection) as the unmodified authorized test. Thus, our SalivaDirect EUA can continue to be modified to fill future needs.

### Limitations of use

Our intended use of SalivaDirect is for the clear and liquid saliva that naturally pools in the mouth. The protocol as currently written is not intended for use of hospitalized COVID-19 patients who are unable to produce “true” saliva. While our previous analysis indicates that saliva is more sensitive for SARS-CoV-2 detection than nasopharyngeal swabs in COVID-19 inpatients (Wyllie et al., 2020), saliva from patients can contain blood or mucus, which can interfere with PCR or make it difficult to pipet. We can overcome many of these issues by having specific collection procedures for saliva collection and using proteinase K to make samples easier to pipet, but some samples can still be invalid if they are not collected properly. While we show that invalid samples are rare from our NBA cohort, our study was biased towards adult men. Saliva collection techniques for adolescents and children, in particular, still need to be refined.

Despite the simplified protocol, SalivaDirect is not a rapid point-of-care test and still requires a complex laboratory. Our method requires electricity and specialized RT-qPCR instruments which can be a limiting factor when such equipment is not available, especially in low- and middle-income countries.

### Target populations and future improvements

To date, there is only one SARS-CoV-2 laboratory diagnostic test authorized by the FDA for asymptomatic testing (U.S. Food & Drug Administration, 2020d), and additional authorized tests are critically needed. While our current EUA is for testing of suspected COVID-19 cases, we designed SalivaDirect to facilitate screening of mostly uninfected populations. Through our partnerships with the NBA and NBPA, we conducted the largest evaluation of saliva to swabs (AN/OP combined) for asymptomatic/presymptomatic SARS-CoV-2 detection. We will use this study to extend our current EUA for asymptomatic screening. Furthermore, we are currently working with testing facilities to evaluate liquid handling robots to decrease sample processing time, and will seek FDA bridging studies to include them on our EUA. Finally, we are initiating preliminary studies to evaluate saliva collection and SARS-CoV-2 detection from school-aged children in Connecticut, US. These key improvements and validation steps will help expand access to testing to help conduct safe in-person learning.

By using many different vendors, not seeking commercialization, and making the protocol completely open, our goal is to make SalivaDirect as accessible as possible. We encourage other groups to make their own adjustments to fit their specific needs or to improve capacity. Thus, our broad FDA EUA application provides a basis for organizations looking to use non-invasive sampling coupled with a simplified molecular testing scheme for SARS-CoV-2 surveillance.

## Methods

### Ethics

The collection of clinical samples from COVID-19 inpatients and healthcare workers at the Yale-New Haven Hospital was approved by the Institutional Review Board of the Yale Human Research Protection Program (Protocol ID. 2000027690). Informed consent was obtained from all patients and healthcare workers prior to sample collection. We used deidentified saliva specimens collected pre-COVID-19 to test for possible cross-reactivity of SalivaDirect. The collection of these saliva specimens was approved by the Institutional Review Board of the Yale Human Research Protection Program (Protocol ID. 0409027018). The collection of deidentified specimens from healthy or asymptomatic individuals from the NBA was approved by the Institutional Review Board of the Yale Human Research Protection Program (Protocol ID. 2000028394). Study participants were informed in writing about the purpose and procedure of the study, and consented to study participation through the act of providing the saliva sample; the requirement for written informed consent was waived by the Institutional Review Board.

### Clinical specimens

Clinical samples were collected from COVID-19 diagnosed patients and healthcare workers at the Yale-New Haven Hospital as described earlier (Vogels et al., 2020b; Wyllie et al., 2020). Briefly, nasopharyngeal swabs were collected in viral transport medium, and saliva was collected in containers without the addition of stabilizing reagents. All specimens were aliquoted upon arrival in the laboratory, with nucleic acid extracted from one aliquot (Ott et al., 2020a), tested using a modified CDC RT-qPCR assay (Vogels et al., 2020b), and the remainder stored at -80°C. We modified the CDC assay by using the 2019-nCoV_N1 (N1), 2019-nCoV-N2 (N2), and human RNase P (RP) primer-probe sets (500 nM of forward and reverse primer and 250 nM of probe per reaction; Integrated DNA Technologies, Coralville, IA, US) with the Luna Universal Probe One-Step RT-qPCR Kit (New England Biolabs, Ipswich, MA, US). Thermocycler conditions were reverse transcription for 10 minutes at 55°C, initial denaturation for 1 min at 95°C, followed by 45 cycles of 10 seconds at 95°C and 30 seconds at 55°C on the CFX96 qPCR machine (Bio-Rad, Hercules, CA, US).

### SalivaDirect protocol

A detailed SalivaDirect protocol has been published (Vogels et al., 2020a). SalivaDirect has been validated with proteinase K and RT-qPCR kits from three vendors, as well as three RT-qPCR instruments (**Table 5**). At least 500 μL of saliva that naturally pools in the mouth was collected in tubes without preservatives. A total of 2.5 μL (50 mg/mL) or 6.5 μL (20 mg/mL) of Proteinase K was added to 50 μL of saliva in 8-strip tubes. The tubes were placed in a rack and vortexed for 1 minute at 3200 RPM. Samples were heated for 5 minutes at 95°C on a thermocycler, and then 5 μL of processed saliva was used as input for the dualplex RT-qPCR assay. The dualplex RT-qPCR assay includes the 2019-nCoV_N1 (N1) primer-probe set that targets the nucleocapsid (N1-F:

GACCC CAAAAT CAG CGAAAT, N1-R: TCTGGTTACTGCCAGTTGAATCTG, N1-P: FAM-ACCCCGCATTACGTTTGGTGGACC-IBFQ) and the human RNase P control (RP) primer-probe set (RP-F: AGATTTGGACCTGCGAGCG, RP-R: GAGCGGCTGTCTCCACAAGT, RP-P: Cy5-TTCTGACCTGAAGGCTCTGCGCG-IBRQ) developed by the CDC (Integrated DNA Technologies, Coralville, IA, US). The fluorophore on the human RNase P probe was modified to combine both primer-probe sets in a dualplex assay, reducing the number of tests to a single assay. Additional fluorophores on the human RNase P probe have been validated and include ATTO647 and Quasar670. Each of these fluorophores are detected by the Cy5 channel (**Table 5**).

In the initial development, we included N2 (Fwd: TTACAAACATTGGCCGCAAA, Rev: GCGCGACATTCCGAAGAA, Probe: HEX-ACAATTTGCCCCCAGCGCTTCAG-IBFQ) (Lu et al., 2020), E (Fwd: ACAGGTACGTT AATAGTTAATAGCGT, Rev: AT ATT GCAGCAGTACGCACACA, Probe: HEX-ACACTAGCCATC CTTACTGCG CTT CG-IBFQ) (Corman et al., 2020), or ORF1 (Fwd: TGGGGYTTTACRGGTAACCT, Rev: AACRCG CTTAACAAAG CACT C, Probe: HEX-TAGTTGTGATGCWATCATGACTAG-IBFQ) (Chu et al., 2020) as a second virus target with HEX-fluorophore. However, this second virus target was removed from the final assay, because unlike the promising results with extracted nucleic acid (Kudo et al., 2020), we were not able to consistently detect SARS-CoV-2 in saliva treated with proteinase K and heat. Thus, the final SalivaDirect dualplex RT-qPCR assay consisted of the N1 and RP primer-probe sets.

The RT-qPCR master mix was prepared following the vendor’s recommended instructions, with 400 nM of N1 forward and reverse primer, 200 nM of N1 probe, 150 nM of RP forward and reverse primer, and 200 nM of RP probe per reaction. Thermocycler conditions were unified for all three RT-qPCR kits (universal protocol) with 10 minutes at 52°C, 2 minutes at 95°C, and 45 cycles of 10 seconds at 95°C and 30 seconds at 55°C. Specimens were considered positive if N1 Ct <40 (or <37 on the ABI 7500 Fast Dx) and any value for RP, negative if N1 Ct ≥40 (or ≥37 on the ABI 7500 Fast Dx) and RP <35, and invalid if N1 Ct ≥40 and RP ≥35. Invalid samples should be retested on a new aliquot of saliva re-run through the entire SalivaDirect protocol.

### Limit of detection

We spiked a positive saliva specimen from a confirmed COVID-19 patient with a known virus concentration (3.7 x 10^4^ copies/μL) into saliva collected from 25 healthcare workers who tested negative for SARS-CoV-2 using the modified CDC assay (Vogels et al., 2020b). We tested a 2-fold dilution series of 400, 200, 100, 50, 25, 12, and 6 SARS-CoV-2 copies/μL in triplicate to determine the preliminary limit of detections, and confirmed the final limit of detection with 20 additional replicates. We used this approach to determine the lower limit of detection of different proteinases K, RT-qPCR kits, and RT-qPCR instruments from multiple vendors (**Table 1**), by using the same input volumes, matrices and RT-qPCR programs for each combination of reagents and instruments. We found no differences in the limit of detection between proteinase K from three vendors and therefore selected one (ThermoFisher Scientific MagMAX proteinase K) to validate the three RT-qPCR kits. The RT-qPCR kit (ThermoFisher TaqPath) with the weakest limit of detection was then used to validate additional RT-qPCR instruments.

### Stability

We determined the stability of SARS-CoV-2 RNA detection in spiked-in saliva samples (12, 25, and 50 copies/μL; as prepared for the limit of detection experiment) by placing them for 7 days at 4°C, room temperature (RT, ~19°C), or 30°C. Results were compared to results obtained in the limit of detection experiment (fresh). Saliva specimens were tested in triplicate and were treated with ThermoFisher Scientific proteinase K and tested with the ThermoFisher TaqPath RT-qPCR kit on the Bio-Rad CFX96.

### Cross-reactivity

We tested 52 saliva specimens, collected from adults during the 2018/2019 and 2019/2020 (pre-COVID19) autumn/winter influenza seasons in New Haven, CT to test for possible cross-reactivity of SalivaDirect with other human respiratory pathogens. Saliva specimens were treated with ThermoFisher Scientific proteinase K and tested with the NEB Luna Universal Probe One-Step RT-qPCR kit on the Bio-Rad CFX96.

### Clinical validation

Paired nasopharyngeal swabs and saliva specimens (collected maximum 4 days apart) were selected from the Yale IMPACT biorepository. In total 67 paired nasopharyngeal swabs and saliva specimens were tested with the US FDA EUA ThermoFisher Scientific TaqPath COVID-19 combo kit following the vendor’s protocol. Briefly, nucleic acid was extracted using the MagMAX Viral/Pathogen Nucleic Acid Isolation Kit on the KingFisher Flex Magnetic Particle Processor. In total 200 μL of specimen was used as input and eluted in 50 μL. For each reaction, 5 μL of extracted nucleic acid was used as input and tested with the ThermoFisher Scientific TaqPath RT-qPCR reaction on the ABI 7500 Fast Dx. Ct values were exported through the 7500 Fast System SDS software v1.4.1. For saliva specimens that were too thick to pipette, 100 μL sample was mixed with 100 μL PBS, and 10 μL was used in the RT-qPCR reaction. For the clinical validation of SalivaDirect, saliva samples were treated with ThermoFisher Scientific proteinase K and tested with the ThermoFisher Scientific TaqPath RT-qPCR kit on the Bio-Rad CFX96.

### Asymptomatic testing

Paired AN/OP swabs and saliva specimens (collected maximum 4 days apart) were collected from NBA players, staff, and other vendors. Combined AN/OP swabs were collected by Quest and followed their EUA specimen collection guidelines (Quest Diagnostics, 2020; U.S. Food & Drug Administration, 2020e). Saliva was collected, under observation of Drug Free Sport International collectors, using a Salimetrics Saliva Collection Aid (Salimetrics, State College, PA, US) into a 2 mL screw-top tube with O-ring cap (Millipore Sigma, Burlingston, MA, US). Saliva was shipped overnight in NanoCool cooling system boxes to our laboratory at the Yale School of Public Health for testing.

A total of 3,779 paired samples were tested by SalivaDirect for saliva specimens at Yale and with Quest’s protocol for swabs, in their labs during the in-market phase of the study while swab testing was completed by BioReference while the teams were in Orlando, FL. Additional swab testing was performed in our lab using the modified CDC multiplex assay protocol for positive swabs sent on for sequencing (Kudo et al., 2020). In total 10 matched samples, reported positive by the NBA, were tested in-house.

### Statistical analysis

GraphPad Prism 8.3.0 was used to make the figures and perform all statistical analyses. Kruskal-Wallis tests were used to test for statistical differences in SARS-CoV-2 RNA stability kept at different temperatures and multiple comparisons were corrected with Dunn’s test. The Wilcoxon matched pairs test was used to test for statistical differences between paired samples. If a virus target was not detected, the Ct value was set to 45 Ct. In all statistical tests, *P* ≤ 0.05 was considered significant.

## Data Availability

All data are included in this article, the supplementary files, and the Source Data.

## Data availability

All data are included in this article, the supplementary files, and the Source Data.

## Acknowledgements

We are honored to have been supported by the NBA, NBPA, and the Yale community, who shared our vision to democratize SARS-CoV-2 testing. Among them, we gratefully acknowledge David Weiss, Anjali Salooja, Peter Meisel, and Wesley Harris from the NBA who helped to design and coordinate the study, and the study participants from the NBA and the Yale-New Haven Hospital for their time and commitment to the efforts. We also thank all members of the collection teams from Drug Free Sport International and the clinical teams at Yale-New Haven Hospital for their dedication and work which made this study possible; and to the US FDA and many staff and faculty at Yale University for helping to bring SalivaDirect to life. We also thank the NBA testing partners, Quest and BioReference, for their continued willingness to support our research. Finally, we are appreciative of the advice and support from the COVID-19 Sports and Society Working Group, and for the moral support from our friends and family (particularly V. Parsons, S. Taylor, and P. Jack).

## Yale IMPACT Research Team Authors

In alphabetical order:

Kelly Anastasio, Michael H. Askenase, Maria Batsu, Sean Bickerton, Kristina Brower, Molly L. Bucklin, Staci Cahill, Yiyun Cao, Edward Courchaine, Giuseppe DeIuliis, Rebecca Earnest, Bertie Geng, Benjamin Goldman-Israelow, Ryan Handoko, William Khoury-Hanold, Daniel Kim, Lynda Knaggs, Maxine Kuang, Sarah Lapidus, Joseph Lim, Melissa Linehan, Alice Lu-Culligan, Anjelica Martin, Irene Matos, David McDonald, Maksym Minasyan, Maura Nakahata, Nida Naushad, Jessica Nouws, Abeer Obaid, Camila Odio, Ji Eun Oh, Saad Omer, Annsea Park, Hong-Jai Park, Xiaohua Peng, Mary Petrone, Sarah Prophet, Tyler Rice, Kadi-Ann Rose, Lorenzo Sewanan, Lokesh Sharma, Albert C. Shaw, Denise Shepard, Mikhail Smolgovsky, Nicole Sonnert, Yvette Strong, Codruta Todeasa, Jordan Valdez, Sofia Velazquez, Pavithra Vijayakumar, Elizabeth B. White, Yexin Yang.

## Author contributions

Designed the laboratory experiments: CBFV, DEB, ALW, and NDG.

Performed the experiments: CBFV, AEW, CAH, DEB, JW, CCK, JRF, and IMO.

Provided clinical samples: JS, EK, PL, AV, MT, AJM, MCM, AC-M, JF, SB, MC, RD, AN, Yale IMPACT Research Team, CSDC, AIK, AI, JM, SFF, and RS.

Designed the NBA study: JS, CC, HMK, JM, SFF, RS, ALW, and NDG.

Analyzed the data: CBFV, AEW, CAH, JW, and JRF.

Supervised the project: CSDC, AIK, AI, JM, PH, CL, SFF, RS, ALW, and NDG.

Wrote and edited the manuscript: CBFV, AEW, CAH, DEB, ALW, and NDG.

All authors read and approved the final manuscript.

## Supplementary figures

**Supplementary Fig. 1:**
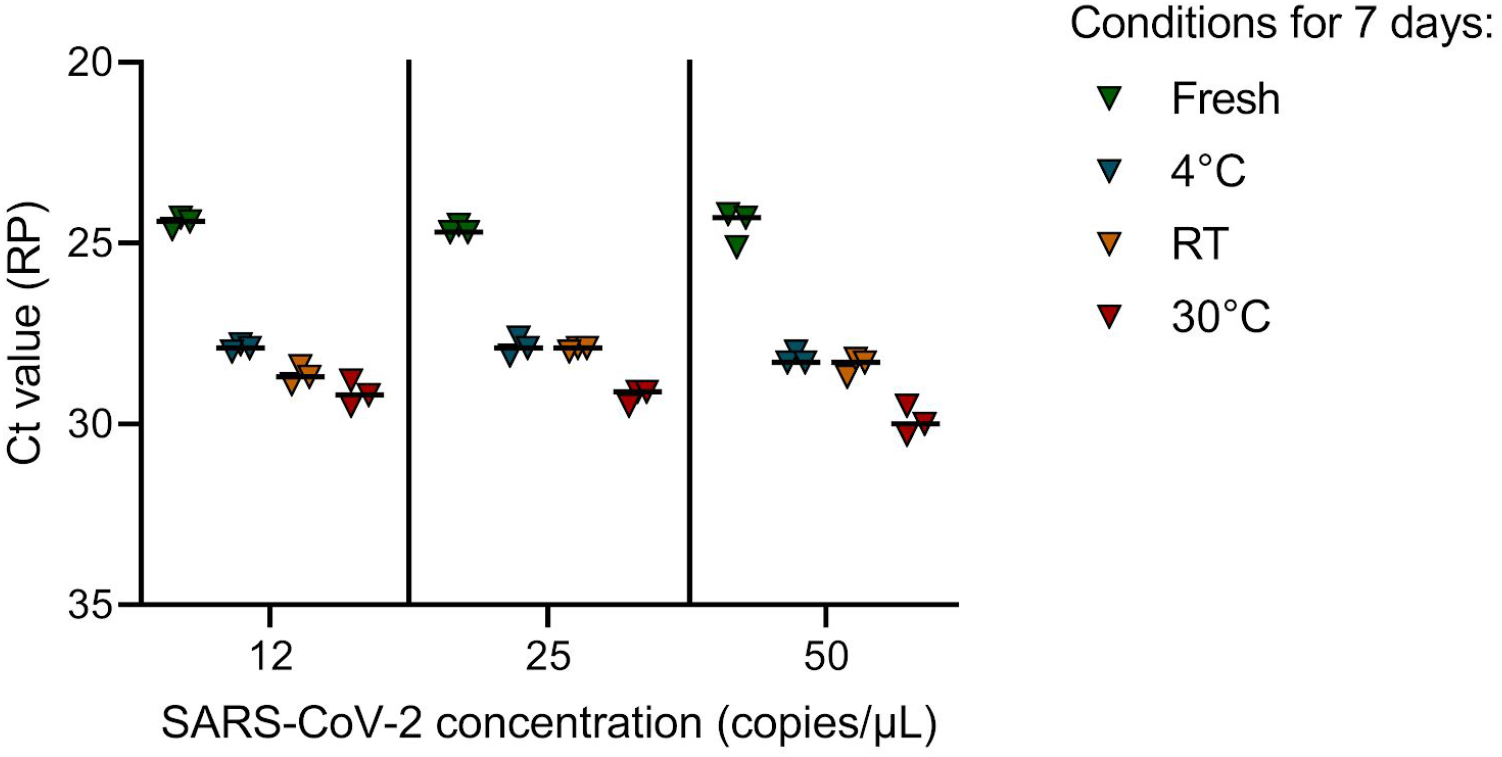
Human RNA in saliva specimens degrades when stored unfrozen for 7 days. When saliva specimens with 12, 25, and 50 SARS-CoV-2 copies/μL were stored under different temperatures for 7 days, we found a significant increase in Ct values for human RNAse P (RP) at RT (Kruskal-Wallis; *P <* 0.01) and 30°C (Kruskal-Wallis; *P* < 0.001), while SARS-CoV-2 N1 Ct values were significantly decreased after 7 days at 30°C (Kruskal-Wallis, *P* = 0.03). This suggests that SARS-CoV-2 is stable in saliva, whereas human RNA seems to degrade over time. The horizontal bars indicate the median. Data used to make this figure can be found in **Source Data Supplementary Fig. 1**.

**Supplementary Fig. 2:**
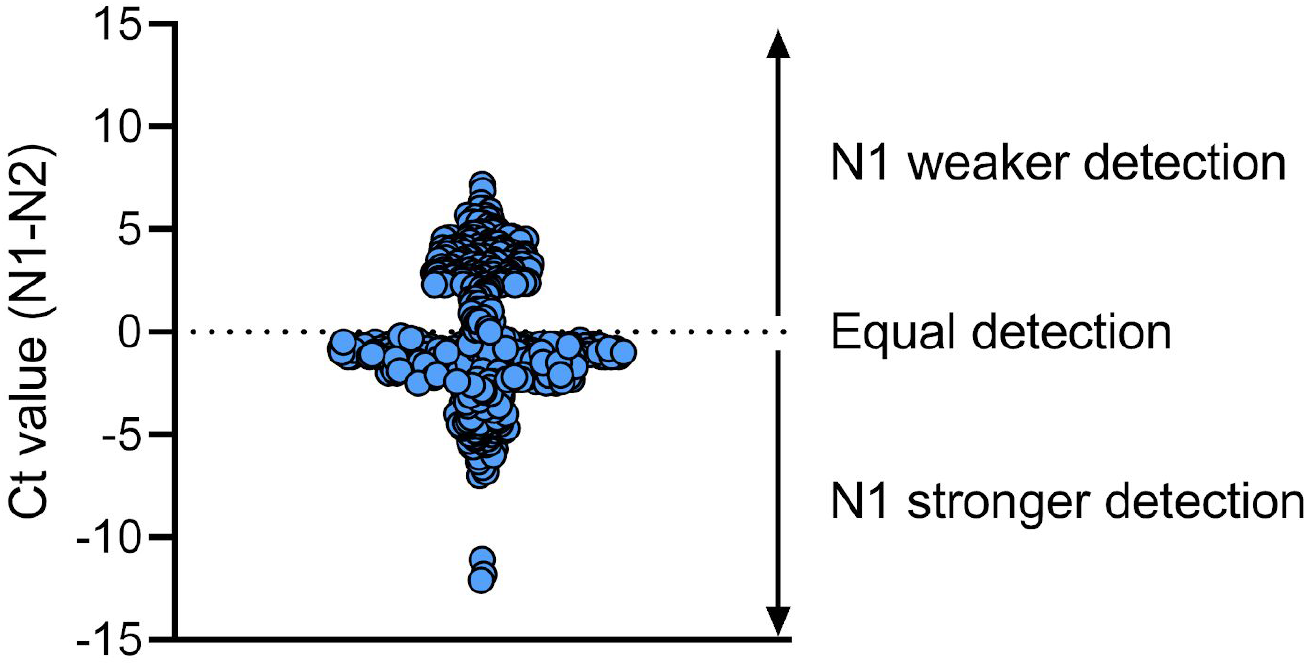
The N1 primer-probe set is more reliable than N2 for SARS-CoV-2 detection. We compared Ct values for N1 and N2 primer-probe sets for 613 clinical specimens, and found that overall the N1 primer-probe set detects a stronger signal as compared to N2. Shown is the difference in Ct value between N1 and N2 and the dotted line indicates equal Ct values for N1 and N2. Data used to make this figure can be found in **Source Data Supplementary Fig. 2**.

**Supplementary Fig. 3:**
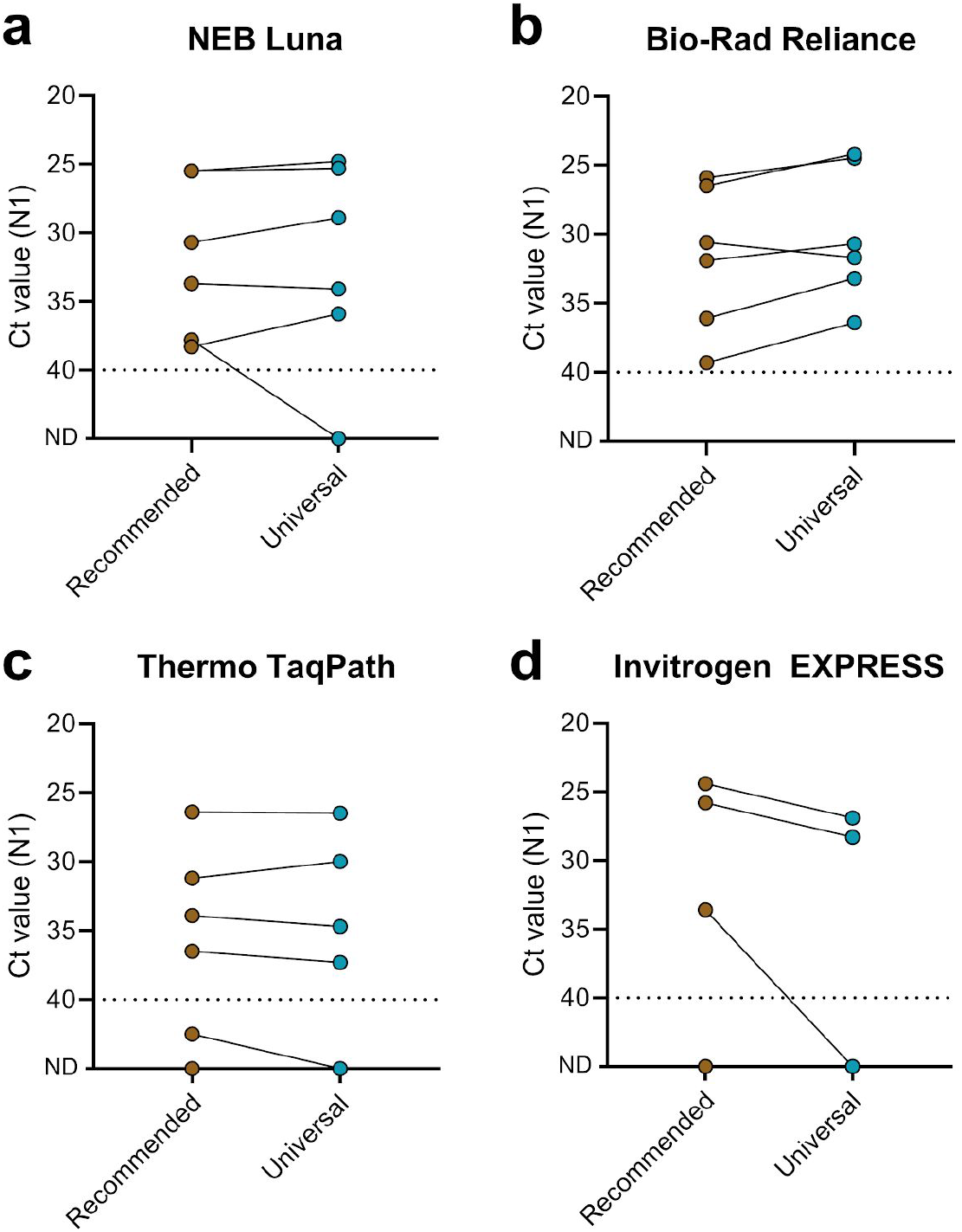
SalivaDirect can be run with RT-qPCR reagents from multiple vendors with universal thermocycler conditions. We selected six positive saliva specimens and tested each sample with four different RT-qPCR kits under recommended and unified thermocycler conditions. The tested kits included the (**a**) NEB Luna Universal Probe One-Step RT-qPCR Kit, (**b**) Bio-Rad Reliance One-Step Multiplex RT-qPCR Supermix, (**c**) TaqPath 1-Step RT-qPCR Master Mix, GC, and (**d**) Invitrogen EXPRESS One-Step superscript qRT-PCR kit. Overall, modifying the thermocycler conditions did not affect the Ct values generated with the N1 primer-probe set (Wilcoxon; Luna: P=0.69, Reliance: P=0.06, TaqPath: P=0.44, EXPRESS: P=0.25). One out of the four evaluated RT-qPCR kits *(e.g*. Invitrogen EXPRESS) was not suitable for SARS-CoV-2 detection with SalivaDirect and was therefore not included in further validation. Shown are the Ct values for the N1 primer-probe set and the dotted line indicates the limit of detection. Data used to make this figure can be found in **Source Data Supplementary Fig. 3**.

**Supplemental Fig. 4:**
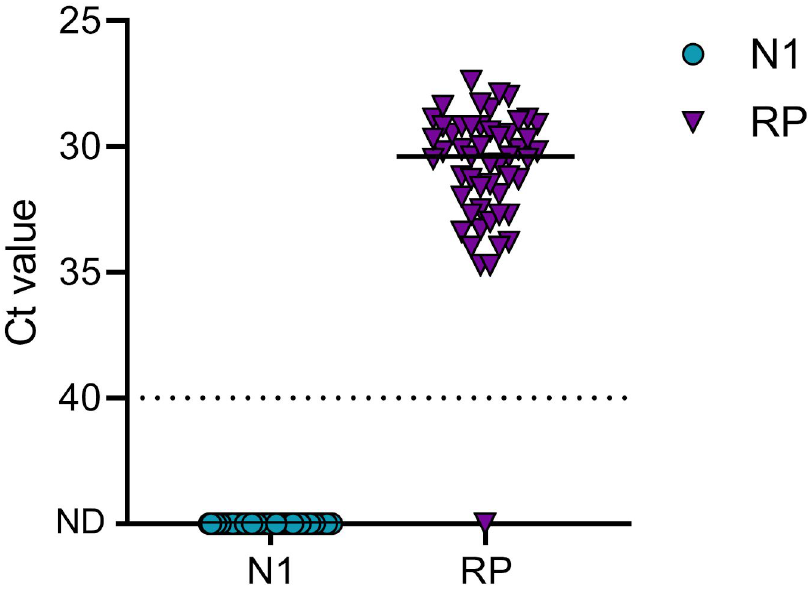
No background amplification when testing pre-COVID-19 saliva specimens with SalivaDirect. Saliva specimens were collected from adults during the 2018-2019 and 2019-2020 autumn/winter influenza seasons and tested with SalivaDirect. Shown are Ct values for N1 and the human RNase P (RP) primer-probe sets. All samples tested negative for N1, indicating no cross-reactivity, while detection of RP indicated proper sample processing. One specimen tested invalid with no detection for both N1 and RP. Shown are the Ct values for the N1 and RP (specimen quality control) primer-probe sets. The horizontal bar indicates the median and the dotted line indicates the limit of detection. Data used to make this figure can be found in **Source Data Supplementary Fig. 4**.

**Supplemental Fig. 5:**
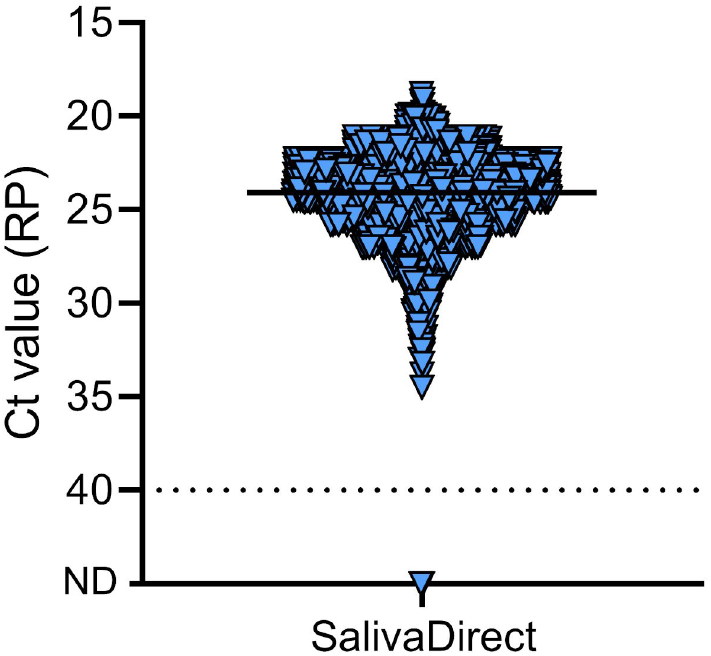
SalivaDirect yields valid results for 99.7% of tested saliva specimens from asymptomatic individuals. A total of 3,779 saliva specimens were collected from asymptomatic NBA players, staff, and contractors and tested with SalivaDirect. After initial testing 38 specimens tested invalid (RP > 35), and after retesting 12 specimens (0.3%) remained invalid. Shown are final human *RNase P values*. The horizontal bar indicates the median and the dotted line indicates the limit of detection. Data used to make this figure can be found in **Source Data Supplementary Fig. 5**.

**Supplementary Fig. 6:**
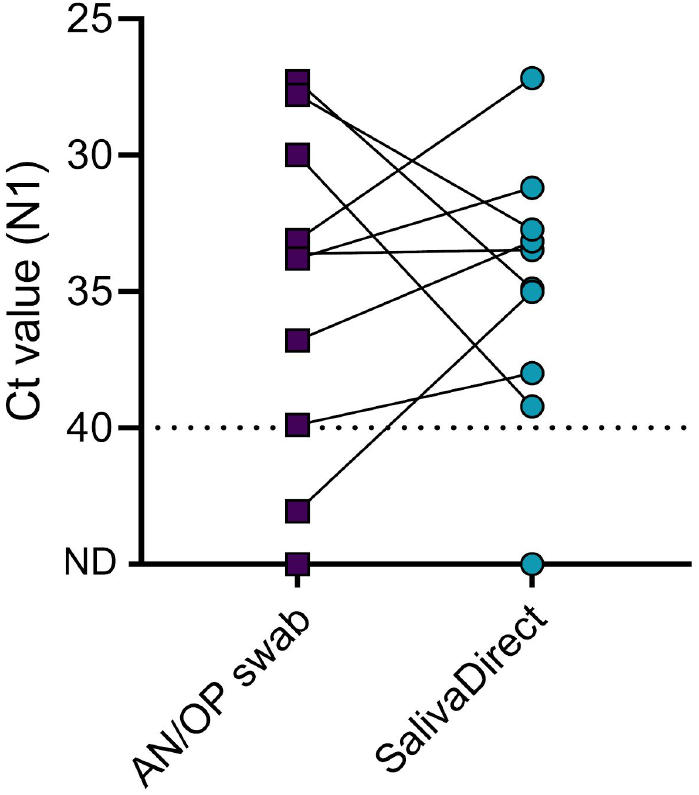
SalivaDirect is comparable for SARS-CoV-2 detection in asymptomatic individuals. We received ten paired AN/OP swabs which were identified as positive when tested by Quest/Bioreference. All ten swabs were retested with the modified multiplex CDC assay and Ct values were compared to saliva tested with SalivaDirect. No significant differences were found between Ct values of AN/OP swabs and saliva (Wilcoxon, *P* = 0.91). Upon retesting, two AN/OP swabs tested negative, of which the paired saliva of one also tested negative by SalivaDirect. Shown are the Ct values for the N1 primer-probe set and the dotted line indicates the limit of detection. Data used to make this figure can be found in **Source Data Supplementary Fig. 6**.

## Supplementary tables

**Supplementary Table S1:** No consistent SARS-CoV-2 detection when testing saliva with a multiplex RT-qPCR assay using a HEX-fluorophore. We compared Ct values between the modified CDC assay with 3 versions of a multiplexed assay with N2 (Fwd: TTACAAACATTGGCCGCAAA, Rev: GCGCGACATTCCGAAGAA, Probe: HEX-ACAATTTGCCCCCAGCGCTTCAG-IBFQ) (Lu et al., 2020), E (Fwd: ACAGGTACGTTAATAGTTAATAGCGT, Rev: ATATTGCAGCAGTACGCACACA, Probe: HEX-ACACTAGCCATCCTTACTGCGCTTCG-IBFQ) (Corman et al., 2020), or ORF1 (Fwd: TGGGGYTTTACRGGTAACCT, Rev: AACRCGCTTAACAAAGCACTC, Probe: HEX-TAGTTGTGATGCWATCATGACTAG-IBFQ) (Chu et al., 2020) as a second virus target with HEX-fluorophore. Eight samples were tested in duplicate with the modified CDC assay (singleplex) as well as each multiplex assay, and average Ct values are shown. No consistent detection of SARS-CoV-2 was achieved for N2, E, or ORF1 with the HEX-fluorophore.

|  | Singleplex |  |  | Multiplex |  |  |  |  |  |  |  |  |
| --- | --- | --- | --- | --- | --- | --- | --- | --- | --- | --- | --- | --- |
|  | N1-FAM | N2-FAM | RP-FAM | N1-FAM | N2-HEX | RP-Cy5 | N1-FAM | E-HEX | RP-Cy5 | N1-FAM | ORF1-HEX | RP-Cy5 |
| 1 | 33.3 | 34.6 | 25.3 | 31.8 | ND | 21.4 | 31.9 | 32.5 | 21.2 | 32.0 | ND | 21.9 |
| 2 | 33.2 | 34.6 | 21.5 | 33.5 | ND | 19.6 | ND | ND | 20.3 | ND | ND | 21.6 |
| 3 | 37.5 | 37.9 | 25.9 | ND | ND | 20.3 | ND | ND | 20.1 | ND | ND | 20.1 |
| 4 | 30.2 | 33.0 | 18.5 | 30.4 | ND | 19.9 | 29.9 | ND | 19.2 | 29.8 | ND | 19.1 |
| 5 | 30.2 | 32.1 | 22.1 | 33.3 | ND | 21.6 | 31.0 | ND | 19.7 | 31.8 | ND | 20.6 |
| 6 | 29.2 | 30.2 | 26.9 | 23.6 | 26.2 | 22.2 | 23.3 | 23.8 | 20.8 | 23.2 | ND | 20.7 |
| 7 | 35.0 | 35.1 | 23.7 | 40.4 | ND | 22.9 | ND | ND | 22.7 | 35.9 | ND | 22.9 |
| 8 | 40.0 | 43.9 | 19.4 | 38.1 | ND | 19.7 | ND | ND | 19.1 | ND | ND | 19.1 |

